# Markers of immune activation and inflammation in individuals with post-acute sequelae of SARS-CoV-2 infection

**DOI:** 10.1101/2021.07.09.21260287

**Authors:** Michael J. Peluso, Scott Lu, Alex F. Tang, Matthew S. Durstenfeld, Hsi-en Ho, Sarah A. Goldberg, Carrie A. Forman, Sadie E. Munter, Rebecca Hoh, Viva Tai, Ahmed Chenna, Brandon C. Yee, John W. Winslow, Christos J. Petropoulos, Bryan Greenhouse, Peter W. Hunt, Priscilla Y. Hsue, Jeffrey N. Martin, J. Daniel Kelly, David V. Glidden, Steven G. Deeks, Timothy J. Henrich

## Abstract

**BACKGROUND:** The biological processes associated with post-acute sequelae of SARS-CoV-2 infection (PASC) are unknown.

**METHODS:** We measured soluble markers of inflammation in a SARS-CoV-2 recovery cohort at early (<90 days) and late (>90 days) timepoints. We defined PASC as the presence of one or more COVID-19-attributed symptoms beyond 90 days. We compared fold-changes in marker values between those with and without PASC using mixed effects models with terms for PASC and early and late recovery time periods.

**RESULTS:** During early recovery, those who went on to develop PASC generally had higher levels of cytokine biomarkers including TNF-alpha (1.14-fold higher mean ratio, 95%CI 1.01-1.28, p=0.028) and IP-10 (1.28-fold higher mean ratio, 95%CI 1.01-1.62, p=0.038). Among those with PASC, there was a trend toward higher IL-6 levels during early recovery (1.28-fold higher mean ratio, 95%CI 0.98- 1.70, p=0.07) which became more pronounced in late recovery (1.44-fold higher mean ratio, 95%CI: 1.11-1.86, p<0.001). These differences were more pronounced among those with a greater number of PASC symptoms.

**CONCLUSIONS:** Persistent immune activation may be associated with ongoing symptoms following COVID-19. Further characterization of these processes might identify therapeutic targets for those experiencing PASC.

## BACKGROUND

The acute phase of SARS-CoV-2 infection is characterized by inflammation and immune dysregulation [1,2], but the recovery phase that follows acute infection is poorly understood. A significant proportion of individuals recovering from COVID-19 do not demonstrate a full return to baseline health and experience post-acute sequelae of SARS-CoV-2 infection (PASC), a condition that is often associated with the persistence or recurrence of symptoms not explained by an alternative medical diagnosis [3]. Multidisciplinary efforts are underway to characterize the epidemiology, natural history, and biology of this condition [4,5], but there is limited biological information regarding its predictors and correlates.

Inflammation during early infection has been associated with adverse outcomes, particularly in those who were hospitalized with COVID-19 [1,2,6–10]. Emerging data suggest that inflammation related to acute SARS-CoV-2 infection can persist for weeks to months [11,12]. For example, individuals well enough to donate convalescent plasma have elevations in certain markers of inflammation, as compared to pre-pandemic plasma donors [11]. These markers include interferon (IFN)-gamma, certain interleukin (IL) proteins, and monocyte chemoattractant protein (MCP)-1. Furthermore, viral proteins and nucleic acids have been detected months after the acute phase in some small studies [13]. However, there are limited data from COVID-19 recovery cohorts with large numbers of individuals managed in the outpatient setting, which constitute the majority of those infected with SARS-CoV-2 and from cohorts with careful prospective clinical phenotyping of symptoms present during COVID-19 recovery.

The clinical implications of persistent inflammation have been established for chronic infections, including HIV [10,11], but are less well understood for acute infections like SARS-CoV-2. We therefore implemented in April 2020 a prospective cohort study of individuals who had documented SARS-CoV-2 infection (Long-term Impact of Infection with Novel Coronavirus, LIINC, www.liincstudy.org; NCT04362150). The majority of participants had not been hospitalized, and biologic specimens and detailed clinical data were collected at regular time points. Here, we report on clinical data and markers of systemic immune activation and inflammation that may contribute to the early understanding of PASC pathogenesis. A better understanding of these pathogenic processes is critical for identifying therapies to treat and/or prevent this condition among the millions of individuals who have recovered from acute SARS-CoV-2 infection.

## METHODS

### Study participants and procedures

Volunteers with a documented history of SARS-CoV-2 infection confirmed by nucleic acid amplification testing were enrolled at our clinical research center at San Francisco General Hospital in San Francisco, California [14]. At each study visit, participants were systematically queried regarding the presence of 32 individual symptoms derived from the Centers for Disease Control list of COVID- 19 symptoms [15] and from the Patient Health Questionnaire Somatic Symptom Scale [16]. A symptom was considered to be present if it was new in onset since the time of SARS-CoV-2 infection or, for pre-existing symptoms, if it had worsened since the diagnosis of SARS-CoV-2 infection. Symptoms that had been present prior to SARS-CoV-2 infection and did not worsen were not considered to represent PASC. At each visit, blood was collected by venipuncture. Plasma was isolated via centrifugation of heparinized blood and stored at −80°C. Due to the known associations between HIV infection and chronic inflammation [10,11], we excluded people living with HIV infection from the current analyses.

### Clinical measurements

We assessed the presence or absence of symptoms at a visit occurring more than 90 days from initial COVID-19 symptom onset. The primary outcome (PASC) was defined as the presence of one or more symptoms at this timepoint. A subset of participants (92/121, 76%) had an additional blood sample that had been collected at an earlier (i.e., before 90 days) timepoint. For further sensitivity and exploratory analyses we defined the top 25% most symptomatic individuals (based on the number of symptoms reported) as having severe PASC.

### Biomarker and antibody assays

The fully automated HD-X Simoa platform was used to measure biomarkers in blood plasma including monocyte chemoattractant protein 1 (MCP-1), Cytokine 3-PlexA (IL-6, IL-10, TNFa), interferon gamma-induced protein-10 (IP-10), interferon-gamma (IFNg), and SARS-CoV-2 receptor binding domain (RBD) IgG according to the manufacturer’s instructions. These markers were selected based upon their biological importance during acute SARS-CoV-2 infection [1,2]. Samples were assayed blinded with respect to associated patient and clinical information. Assay performance was consistent with the manufacturer’s specifications.

### Statistical analysis

We used descriptive statistics to characterize the cohort and non-parametric analyses to compare the groups with and without PASC. We log10-transformed all biomarkers as they were not normally distributed. We then compared the ratio of the mean transformed values for each biomarker between those with and without persistent symptoms using mixed effects models with terms for PASC and time period (early versus late recovery). This approach permits comparison of the values at early and late time points as well as assessment of whether trajectories in marker values differ between those with and without persistent symptoms. We calculated fold-changes and 95% confidence intervals by exponentiating the coefficients to give the ratio between the untransformed biomarker values. We used Spearman correlations to evaluate relationships between levels of binding antibodies and immune markers. All p-values are two sided. We used Stata (version 16.1; StataCorp, College Station, TX) and Prism (version 9.1.2, GraphPad Software, L.L.C., San Diego, CA).

### Ethics approval

All participants provided written informed consent. The study was approved by the Institutional Review Board at the University of California, San Francisco.

## RESULTS

### Study participants

Our analysis cohort included 121 participants and was diverse in terms of sex, gender, race, ethnicity, and level of education (Table 1). Medical comorbidities present in more than 10% of the cohort included lung problems, diabetes, and obesity. Nine participants had a history of an autoimmune disorder, which included autoimmune thyroid disease (n=4), gastrointestinal disease (n=2), multiple sclerosis (n=1), rheumatoid arthritis (n=1), and an unspecified autoimmune condition (n=1). A majority of participants (78%) had been managed as outpatients during their COVID-19 illness.

**Table 1.** Characteristics of study cohort. IQR, interquartile range. P-values represent either Mann-Whitney U tests for continuous variables or chi-square test for categorical variables. Fisher’s exact test was used when individual cell counts were for categorical variables.

| Characteristic | All<br>(n=121) | No PASC<br>(n=48) | PASC<br>(n=73) | P-Value<br>(PASC vs no PASC) |
| --- | --- | --- | --- | --- |
| **Demographic characteristics** |  |  |  |  |
| **Age (IQR)** | 44 (37-57) | 44.5 (37-58.5) | 44 (36-56) | P=0.78 |
| **Sex assigned at birth** |  |  |  | P=0.053 |
| Female | 66 (54.5%) | 21 (43.8%) | 45 (61.6%) |  |
| Male | 55 (45.5%) | 27 (56.3%) | 28 (38.4%) |  |
| **Gender** |  |  |  | P=0.063 |
| Cisgender Female | 65 (53.7%) | 21 (43.8%) | 44 (60.3%) |  |
| Cisgender Male | 54 (44.6%) | 27 (56.3%) | 27 (37.0%) |  |
| Transgender Male | 2 (1.7%) | 0 (0.0%) | 2 (2.7%) |  |
| **Race and ethnicity** |  |  |  | P=0.20 |
| Hispanic/Latino | 30 (24.8%) | 8 (16.7%) | 22 (30.1%) |  |
| White | 69 (57.0%) | 28 (58.3%) | 41 (56.2%) |  |
| Black/African American | 3 (2.5%) | 1 (2.1%) | 2 (2.7%) |  |
| Asian | 14 (11.6%) | 8 (16.7%) | 6 (8.2%) |  |
| Pacific Islander/Native Hawaiian | 1 (0.8%) | 1 (2.1%) | 0 (0.0%) |  |
| Not Provided | 4 (3.3%) | 2 (4.2%) | 2 (2.7%) |  |
| **Highest level of education completed** |  |  |  | P=0.46 |
| At least some high school | 21 (17.4%) | 9 (18.8%) | 12 (16.4%) |  |
| At least some college | 46 (38.0%) | 15 (31.3%) | 31 (42.5%) |  |
| At least some graduate school | 54 (44.6%) | 24 (50.0%) | 30 (41.1%) |  |
| **Tobacco use history** | 27 (22.3%) | 9 (18.8%) | 18 (24.7%) | P=0.45 |
| **Clinical characteristics** |  |  |  |  |
| **Pre-existing medical conditions** |  |  |  |  |
| Autoimmune disease | 9 (7.4%) | 1 (2.1%) | 8 (11.0%) | P=0.085 |
| Cancer (with treatment received within 2 years prior to COVID-19 diagnosis) | 3 (2.5%) | 1 (2.1%) | 2 (2.7%) | P=0.65 |
| Diabetes | 14 (11.6%) | 6 (12.5%) | 8 (11.0%) | P=0.82 |
| Lung problems | 23 (19.0%) | 10 (20.8%) | 13 (17.8%) |  |
| **Body mass index category** |  |  |  | P=0.20 |
| 24.9 or less | 46 (38.0%) | 20 (41.7%) | 26 (35.6%) |  |
| 25 to 29.9 | 34 (28.1%) | 16 (33.3%) | 18 (24.7%) |  |
| 30 or greater | 39 (32.2%) | 11 (22.9%) | 28 (38.4%) |  |
| **Hospitalized during acute COVID-19** | 27 (22.3%) | 8 (16.7%) | 19 (26.0%) | P=0.23 |

Although we did not identify significant demographic differences between groups, those reporting PASC tended to be more likely to have been assigned female sex at birth (61.6% vs 43.8%, p=0.053) and to report a history of autoimmune disease preceding their COVID-19 diagnosis (11% vs 2.1%, p=0.087). The presence of PASC did not differ according to hospitalization status during acute COVID-19.

The early recovery measurement occurred at a median of 52 (interquartile range [IQR] 38-64) days from symptom onset and the late recovery measurement occurred at a median of 124 (IQR 116-136) days from symptom onset. The timing of the early follow-up sample among those with PASC was slightly later than those without PASC (median 57 vs 50 days after symptom onset, p=0.04); the timing of the late sample did not differ (median 123 vs 124 days after symptom onset, p=0.26).

### Persistent symptoms

Seventy-three individuals reported one or more symptoms at the late recovery timepoint. The median number of symptoms reported by individuals with PASC was 5 (IQR 2-8, absolute range 1-18). Common symptoms (Table 2) included problems with memory or concentration (57%), fatigue (56%), shortness of breath (38%), and anosmia/dysgeusia (37%).

**Table 2.** Symptoms reported at late follow-up among those with PASC. Participants were systematically asked about 32 individual symptoms at the late follow-up visit, which took place a median of 124 days from initial COVID-19 symptom onset.

| Symptoms reported at late follow-up | N=73<br>n (%) |
| --- | --- |
| Constitutional |  |
| *Fatigue* | 41 (56.2%) |
| *Subjective fever* | 2 (2.7%) |
| *Chills* | 2 (2.7%) |
| *Objective fever* | 2 (2.7%) |
| Upper Respiratory |  |
| *Rhinorrhea* | 14 (19.2%) |
| *Sore throat* | 7 (9.6%) |
| Cardiopulmonary |  |
| *Cough* | 13 (17.8%) |
| *Shortness of breath* | 28 (38.4%) |
| *Chest pain* | 18 (24.7%) |
| *Palpitations* | 15 (20.5%) |
| *Fainting* | 0 (0.0%) |
| Gastrointestinal |  |
| *Diarrhea* | 9 (12.3%) |
| *Nausea* | 17 (23.3%) |
| *Loss of appetite* | 12 (16.4%) |
| *Abdominal pain* | 6 (8.2%) |
| *Vomiting* | 1 (1.4%) |
| *Constipation* | 3 (4.1%) |
| Genitourinary |  |
| *Menstrual cramps* | 2 (2.7%) |
| *Dyspareunia* | 0 (0.0%) |
| Rash | 10 (13.7%) |
| Musculoskeletal |  |
| *Myalgia* | 20 (27.4%) |
| *Back pain* | 7 (9.6%) |
| *Joint pain* | 9 (12.3%) |
| Anosmia/Dysgeusia | 27 (37.0%) |
| Neurologic |  |
| *Headache* | 18 (24.7%) |
| *Concentration problems* | 42 (57.5%) |
| *Dizziness* | 18 (24.7%) |
| *Balance problems* | 13 (17.8%) |
| *Neuropathy* | 17 (23.3%) |
| *Vision problems* | 13 (17.8%) |
| *Parosmia* | 8 (11.0%) |
| Trouble Sleeping | 32 (43.8%) |

### Levels of plasma biomarkers among those with and without PASC

In our cross-sectional analyses, we first compared levels of each plasma marker measured during early recovery between those with and without PASC (Figure 1). Those who went on to develop PASC demonstrated significantly higher levels of TNF-alpha (1.14-fold higher mean ratio, 95% CI 1.01-1.28, p=0.028) and IP-10 (1.28-fold higher mean ratio, 95% CI 1.01-1.62, p=0.038). The mean IL-6 level during early recovery was on average 29% higher among those with PASC, although the difference did not achieve statistical significance (95% CI 0.98-1.70, p=0.07). Several other markers showed higher levels among those with PASC even when the comparisons were not significant.

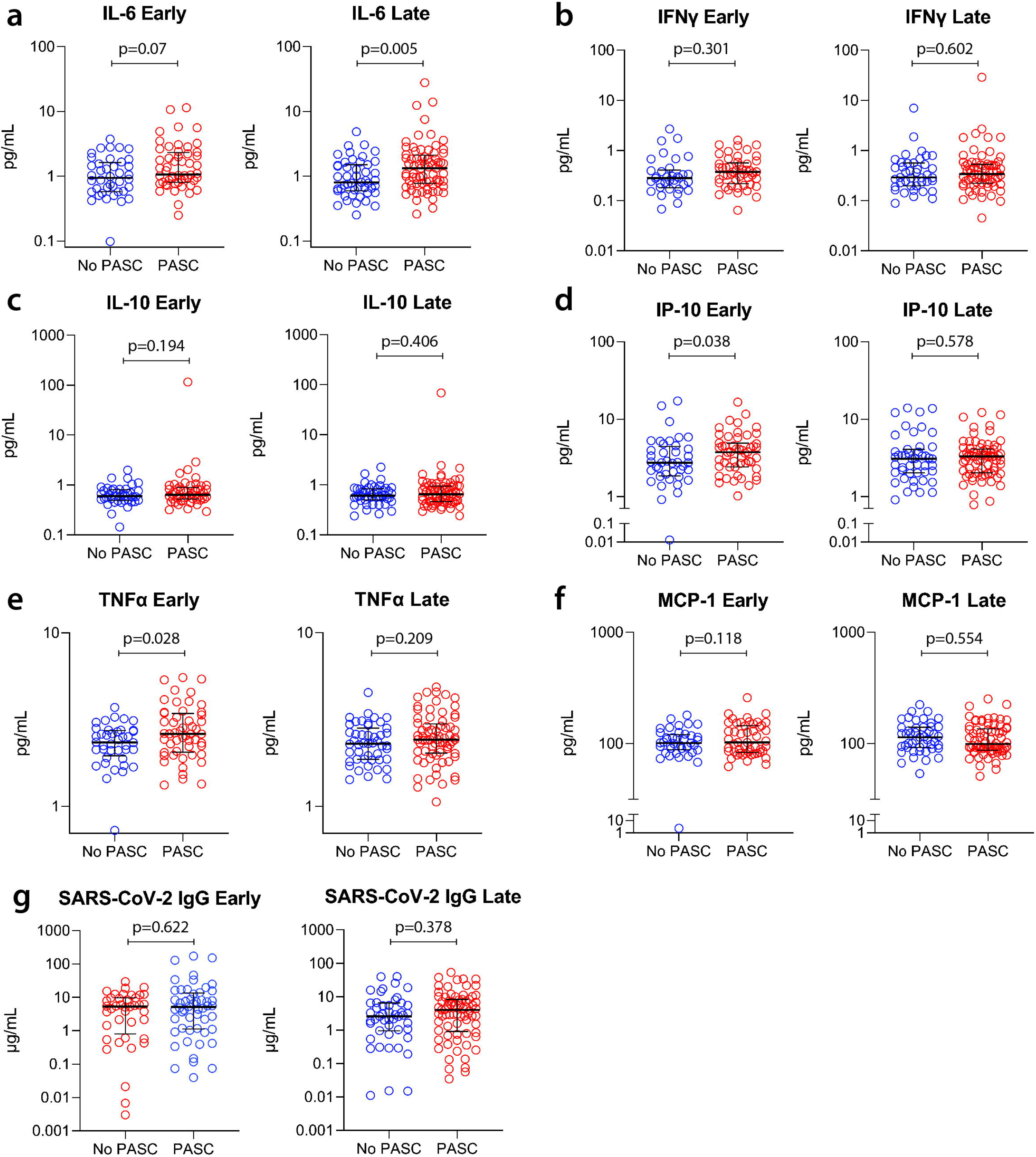

We next compared levels of each biomarker measured during late recovery between those with and without PASC (Figure 1). During the late recovery period, the mean IL-6 level was on average 44% higher among those with PASC (95% CI:1.11-1.86, p=0.0005). No other markers differed between the groups.

Levels of SARS-CoV-2 RBD IgG did not differ between groups at either the early or late timepoints. Among those with PASC, levels of binding antibodies correlated with TNF-alpha (r=0.33, p=0.018), interferon-gamma (r=0.30, p=0.038), and MCP-1 (r=0.39, p=0.005) at the early timepoint; at the later timepoint, levels of binding antibodies correlated with IL-6 (r=0.29, p=0.015), TNF-alpha (r=0.28, r=0.016), and MCP-1 (r=0.40, p=0.0006). In the group without PASC, these antibodies correlated only with interferon-gamma at the early timepoint (r=0.38, p=0.024) and IP-10 at the late timepoint (r=0.33, p=0.025).

### Changes in levels of plasma biomarkers over time

In our longitudinal analyses, we used mixed models to indicate changes in levels of biomarkers among those with and without PASC between the early and late recovery period time points (Figure 2). Overall, there were no statistically significant differences in the trends of biomarkers over time between the PASC and non-PASC groups. As would be predicted from cross-sectional analyses, higher levels of IL-6 were observed across time points in those with PASC compared to those without persistent symptoms.

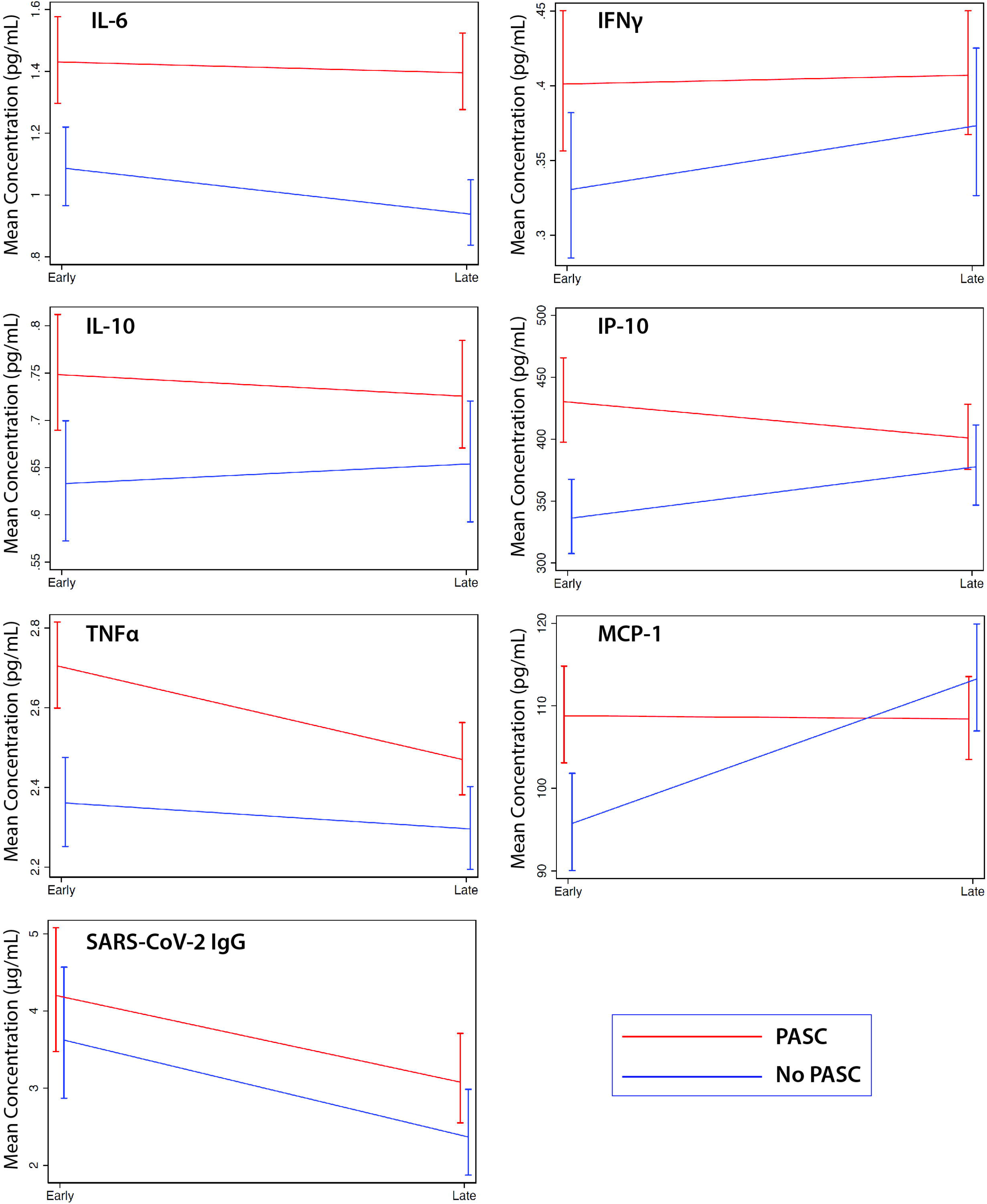

### Sensitivity analyses

To further assess the effects of certain demographic factors potentially important in PASC, we performed adjustment for age, sex, and hospitalization status during acute SARS-CoV-2 infection. These adjustments did not fundamentally change the interpretation of the earlier analyses, although the IP-10 elevation during early recovery was no longer statistically significant (mean ratio 1.24 95%CI 0.98-1.57, p=0.071; see Supplemental Table 1).

To determine whether the relationships observed in the primary analysis would be more pronounced among those with the most severe manifestations of PASC, we examined the subset of individuals reporting the greatest number of symptoms (upper 25% of symptom count) in comparison to those without any persistent symptoms. During early recovery, severe PASC was associated with the presence of elevated levels of IL-6 (mean ratio 1.47 95% CI 1.10-1.97, p=0.009), interferon-gamma (mean ratio 1.35 95% CI 1.00-1.81, p=0.049), IL-10 (mean ratio 1.25 95% CI 1.00-1.57, p=0.05), and TNF-alpha (mean ratio 1.16 95% CI 1.03-1.31, p=0.017). IL-6 levels were persistently elevated during late follow-up, although the relationship did not remain statistically significant (mean ratio 1.47 95% CI 1.10-1.97, p=0.089).

## DISCUSSION

Post-acute sequelae of SARS-CoV-2 infection (PASC) is now recognized as a public health priority [3]. In a variety of cohorts and electronic medical record-based studies [4,5,12,17], approximately 10- 40% of individuals report new symptoms that persist for weeks to months. Risk factors for PASC may include more severe acute infection, advanced age, female sex, and socioecomonic factors [5,17,18]. However, the study of the pathogenesis of PASC is only just beginning. Leveraging a prospective cohort designed to study how SARS-CoV-2 infection affects long-term health, we measured a variety of potential biomarkers in individuals recovering from COVID-19, a substantial proportion of whom reported symptoms beyond 3 months following initial infection. Importantly, we did not prospectively seek to enroll individuals with PASC. We identified several biomarkers – particularly IL-6 and TNF- alpha – that differ during early and late recovery among those who continue to experience symptoms at a median of 124 days following infection. These elevations are part of a consistent pattern suggesting persistent immune activation among those who go on to develop PASC. These observations may inform on biological pathways contributing to PASC and aid in the identification of potential therapeutic strategies.

IL-6 and TNF-alpha are both pro-inflammatory cytokines that contribute to leukocyte recruitment, activation, and differentiation, as well as B cell maturation and the expansion of T helper cell subsets [19,20]. Both have been identified as key factors in the immune response during acute COVID-19 [2,9,10,21,22], although in general IL-6 levels appear to be more predictive of poor outcomes such as respiratory failure and the need for mechanical ventilation [2,8,10,23]. Results from interventions to reduce the inflammatory milieu during acute infection by targeting cytokines, such as IL-6, however, have been mixed [24–29]. While various immune activating cytokines can have protective or homeostatic effects [30–32], their dysregulation may lead to detrimental clinical conditions. For example, IL-6 has been implicated in models of chronic inflammatory diseases [30], and overexpression in mouse models is associated with tissue-specific manifestations such as lung or neurologic disease [33–35]. Similarly, TNF-alpha induces tissue inflammation and endothelial activation and uncontrolled activity of this cytokine underlies various inflammatory diseases that can involve multiple organ systems [36].

Among those recovering from COVID-19, there are limited data on immunologic trends over time and in association with ongoing clinical symptoms. Although our findings need to be supported with larger studies, we found that individuals with persistent symptoms were likely to demonstrate higher levels of markers of inflammation and immune activation during the first 90 days of COVID-19 recovery. While not all markers achieved statistical significance, there was a consistent trend across all markers of interest during this period. Furthermore, we found that elevations in IL-6 persisted into the late recovery period during which we defined the primary clinical outcome (PASC). This observation builds upon prior work identifying persistent elevations in levels of cytokines in COVID-19 convalescent plasma donors [11] and individuals who were previously hospitalized with COVID-19 [12]. In the latter study, previously hospitalized individuals who reported persistent symptoms had higher levels of MCP-1 and platelet derived growth factor (PDGF) during early convalescence, but these differences were no longer detected at 6 months and the study included a lower proportion of participants with persistent symptoms. Our findings suggest that differences in cytokines and chemokines may also be relevant among individuals who were not hospitalized during the acute phase, and that persistent immune activation might be associated with the processes that drive some persistent symptoms.

The factors driving persistent inflammation during COVID-19 recovery have yet to be determined. One possibility is that residual inflammation from the acute phase of infection persists among certain individuals, either those with the most severe illness or in relation to other unknown factors. This could be supported by the fact that most differences in the levels of these markers appear to resolve by the late recovery timepoint. Although antibody levels, which consistently correlate with disease severity [37–46] and have been shown to be predictive of persistent symptoms at 6 months in other cohorts [4], did not differ between the groups, among those with PASC antibody levels did correlate with levels of several inflammatory markers. In addition, IL-6 levels remained elevated throughout the observation period. Many factors could contribute to persistent IL-6 elevations including antigen persistence in tissues, compromised mucosal barrier integrity with increased microbial translocation, and/or autoreactive immunity, or an intrinsic failure of the host immune system to return to baseline homeostasis. Further work will be needed to determine whether our findings represent a persistent inflammatory response that is slower to resolve among those with PASC, or whether there is ongoing pathology that would benefit from intervention during either early or later recovery.

It is notable that 8/9 participants with a history of previously diagnosed autoimmune disorders experienced PASC. It has been theorized that elevated autoantibodies may be present during acute hospitalization with COVID-19 and may persist during convalescence [47,48], although the association with persistent symptoms has yet to be established. Others have observed new diagnoses or exacerbations of clinical entities with autoimmune mechanisms, such as diabetes and thyroiditis, following SARS-CoV-2 infection [49,50]. The relationship between SARS-CoV-2 infection, autoreactive immunity, and the inflammatory milieu that may persist following COVID-19 warrants further investigation, particularly if it can be tied to specific PASC phenotypes.

Strengths of this analysis include the inclusion of a large number of individuals with deeply characterized persistent symptoms and the large proportion who had not been hospitalized during acute infection, which is reflective of the majority of people recovering from COVID-19. Limitations of this study include the use of a convenience sample that may not be representative of the SARS-CoV- 2 epidemic and lack of specimens from the acute infection period. There is currently no widely agreed upon case definition for PASC, and we adopted a broad case definition which might be overly sensitive. We also used a rough temporal cutoff to classify PASC, which may have led to misclassification bias in our results. Still, our sensitivity analyses showed similar findings, suggesting the presence of a true effect. We measured a relatively small number of biomarkers based on hypotheses derived from published signatures during acute infection, but more in depth biomarker characterization may be more revealing. While we performed multiple statistical analyses, our findings were consistent with our prespecified hypotheses. Nonetheless, our observation will help inform on mechanistic pathways that could contribute to PASC and direct future research leading to the identification of potential therapeutic targets.

## Data Availability

Data will be made available when the analysis is complete.

## Acknowledgements

We are grateful to the study participants. We are grateful to the LIINC study participants and to the clinical staff who provided care to these individuals during their acute illness period and during their recovery. We thank Dr. Isabel Rodriguez-Barraquer and Dr. Rachel Rutishauser for their contributions to the LIINC leadership team. We acknowledge current and former LIINC clinical study team members Tamara Abualhsan, Andrea Alvarez, Mireya Arreguin, Emily Fehrman, Monika Deswal, Heather Hartig, Yanel Hernandez, Marian Kerbleski, Lynn Ngo, Ruth Diaz Sanchez, Cassandra Thanh, Leonel Torres, Fatima Ticas, and Meghann Williams; and LIINC laboratory team members Joanna Donatelli, Jill Hakim, Nikita Iyer, Owen Janson, Christopher Nixon, and Keirstinne Turcios. We thank Elnaz Eilkhani for coordination with the Institutional Review Board. We acknowledge the contributions of the UCSF Clinical and Translational Science Unit, Core Immunology Laboratory, and AIDS Specimen Bank.

## FOOTNOTES

### Author Contributions

MJP, HH, JDK, JNM, SGD, and TJH designed the study, which was supported through funding to JDK, JNM, SGD, and TJH. MJP, SEM, RH, VT collected clinical data and biospecimens. Specimens were analyzed by AC, BCY, JWW, and CJP. MJP, SL, AT, MSD, JDK, and DVG performed and/or interpreted the statistical analyses. MJP, SL, AT, HH, CF, JDK, SGD, and TJH drafted the initial manuscript with input from AC, JWW, PWH, PSY, and JNM. All authors edited, reviewed, and approved the final manuscript.

