## Supplementary Materials for "Markers of immune activation and inflammation in individuals with post-acute sequelae of SARS-CoV-2 infection"

**Supplemental Tables**

Primary Analysis – PASC at Late Timepoint (>90 days)

|  | **P-value** | | **Fold Change in Mean 95% CI** | | **P-value** |
| --- | --- | --- | --- | --- | --- |
|  | **Early** | **Late** | **Early** | **Late** | **Change over Time** |
| il6 | 0.07 | 0.005 | 1.29 (0.98-1.70) | 1.44 (1.11-1.86) | 0.373 |
| mcp1 | 0.118 | 0.554 | 1.14 (0.97-1.33) | 0.96 (0.83-1.11) | 0.056 |
| ifngama | 0.301 | 0.602 | 1.16 (0.87-1.55) | 1.07 (0.83-1.39) | 0.604 |
| il10 | 0.194 | 0.406 | 1.16 (0.93-1.44) | 1.10 (0.88-1.36) | 0.377 |
| tnfa | 0.028 | 0.209 | 1.14 (1.01-1.28) | 1.07 (0.96-1.20) | 0.206 |
| ip10 | 0.038 | 0.578 | 1.28 (1.01-1.62) | 1.06 (0.86-1.31) | 0.138 |
| igg | 0.622 | 0.378 | 1.16 (0.65-2.05) | 1.29 (0.73-2.26) | 0.383 |

Adjusted Analysis – Adjusted for age, sex, and hospitalization status

|  | **P-value** | | **Fold Change in Mean 95% CI** | | **P-value** |
| --- | --- | --- | --- | --- | --- |
|  | **Early** | **Late** | **Early** | **Late** | **Change over Time** |
| il6 | 0.132 | 0.008 | 1.22 (0.94-1.58) | 1.38 (1.09-1.76) | 0.296 |
| mcp1 | 0.128 | 0.654 | 1.13 (0.97-1.32) | 0.97 (0.84-1.11) | 0.085 |
| ifngama | 0.234 | 0.47 | 1.19 (0.89-1.60) | 1.10 (0.85-1.44) | 0.615 |
| il10 | 0.242 | 0.479 | 1.14 (0.91-1.43) | 1.08 (0.87-1.35) | 0.377 |
| tnfa | 0.02 | 0.143 | 1.14 (1.02-1.28) | 1.08 (0.97-1.20) | 0.252 |
| ip10 | 0.071 | 0.742 | 1.24 (0.98-1.57) | 1.04 (0.84-1.28) | 0.152 |
| igg | 0.929 | 0.718 | 0.98 (0.59-1.63) | 1.10 (0.66-1.81) | 0.352 |

Severe PASC Analysis – Top 25% of symptom number reported at late timepoint

|  | **P-value** | | **Fold Change in Mean 95% CI** | | **P-value** |
| --- | --- | --- | --- | --- | --- |
| **var** | **Early** | **Late** | **Early** | **Late** | **Change over Time** |
| il6 | 0.009 | 0.089 | 1.47 (1.10-1.97) | 1.25 (0.97-1.62) | 0.211 |
| mcp1 | 0.072 | 0.41 | 1.17 (0.99-1.38) | 0.94 (0.81-1.09) | 0.021 |
| ifngama | 0.049 | 0.928 | 1.35 (1.00-1.81) | 0.99 (0.77-1.28) | 0.049 |
| il10 | 0.05 | 0.431 | 1.25 (1.00-1.57) | 1.09 (0.88-1.35) | 0.035 |
| tnfa | 0.017 | 0.211 | 1.16 (1.03-1.31) | 1.07 (0.96-1.20) | 0.125 |
| ip10 | 0.101 | 0.908 | 1.24 (0.96-1.60) | 0.99 (0.80-1.22) | 0.095 |
| igg | 0.627 | 0.317 | 1.16 (0.65-2.07) | 1.33 (0.76-2.35) | 0.275 |
